# High serum Cholesterol and Triglyceride levels in older adults: associations with sleep and nighttime behavior disorders at baseline and a prediction analysis of incidental cases at 12 months follow-up

**DOI:** 10.1101/2024.06.05.24308529

**Authors:** Asma Hallab, the Alzheimer’s Disease Neuroimaging Initiative

## Abstract

**Introduction:** This study explored the association between dyslipidemia and sleep and nighttime behavior disorders (SNBD) in the elderly.

**Methods:** ADNI population with complete Cholesterol, Triglyceride, SNBD, and neurocognitive data were included. Logistic regression was performed to study the association between dyslipidemia and SNBD at baseline and 12 months. Relevant confounders were adjusted for.

**Results:** Among the 2,216 included cases, 1,045 (47%) were females, and the median age was 73 (IQR: 68, 78). At baseline, 357 (16%) had SNBD, and 327 (18%) at 12 months; 187 were incident cases.

There were more cases of baseline SNBD in the hypertriglyceridemia group than in those without (19% vs. 14%, *p*-value=0.003). Similarly, more follow-up SNBD cases had hypertriglyceridemia at baseline (21% vs. 16%, *p*-value=0.025). SNBD cases at baseline had significantly higher serum Triglyceride levels than those without (132 vs. 118mg/dL, *p*-value<0.001).

Only hypertriglyceridemia was significantly associated with baseline SNBD (crude OR=1.43, 95%*CI*: 1.13,1.80, *p*-value=0.003), even after adjustment for confounding factors (adj.OR=1.36, 95%*CI*: 1.06,1.74, *p*-value=0.016) and (BMI-adj.OR=1.29, 95%*CI*: 1.00,1.66, *p-*value=0.048). None of the dyslipidemia forms did predict incident cases at 12 months.

**Conclusions:** Hypertriglyceridemia, but not hypercholesterolemia, was associated with higher odds of SNBD. None of the dyslipidemia forms predicted incidental SNBD over 12 months.

## 1. Introduction

Sleep disorders represent a large spectrum of symptoms defining an alteration of sleep quality, structure, chronobiology, duration, and associated breathing and movement disorders. (1) Insomnia and sleep obstructive apnea syndrome have a high prevalence worldwide; both are associated with cardiovascular and neuropsychiatric risk factors and affected persons are exposed to higher morbidity and mortality rates. (2–5) The incidence of sleep disorders increases with age. In addition to the physiological decrease in sleep hours during the aging process; neurodegeneration, neuroendocrine, and sleep disorders define a more complex bidirectional association. (6) Old patients with sleeping disorders, particularly insomnia, have higher risks of cognitive decline, and those with cognitive impairment are more susceptible to progressing into dementia. (7–9) Moreover, elderly patients with neurocognitive disorders are at higher risk of experiencing neurodegeneration-related sleep behavior and movement disorders. (10)

The relationship between sleep disorders and metabolic and cardiovascular pathologies is largely reported in the literature. (11, 12) Defined as the association between diabetes, dyslipidemia, hypertonia, and visceral adiposity, the metabolic syndrome is a well-established risk factor for cardiovascular disorders and is related to higher morbidity and mortality rates. (13) Most published studies evaluated sleep disorders quantitatively based on sleep duration or qualitatively depending on the subjective perception of sleep quality. (14–17) Studies on sleep disorders objectified by the study partner of older patients and their association with dyslipidemia are limited. Moreover, there is limited data on whether dyslipidemia might predict prospectively sleep disorders.

The aim of this study was (1) to explore the association between dyslipidemia and sleep and nighttime behavior disorders (SNBD) in the elderly and (2) to evaluate whether dyslipidemia at baseline might predict incidental cases of SNBD over 12 months of follow-up.

## 2. Methods

This manuscript has been prepared and reported according to STROBE guidelines. (18)

### Study population

The studied population is part of the Alzheimer’s Disease Neuroimaging Initiative (ADNI) cohort, from which only cases with complete data required for the current analysis were included. ADNI was initiated by Dr Michael W. Weiner. Study participants are older adults recruited at 59 centers around the United States and Canada, who underwent an observational follow-up where biological, genetic, neuroimaging, and neuropsychiatric information was assessed at several time points. Participants from different phases (ADNI 1, go, 2, and 3) were eligible for the current analysis. Data, ethical approval, enrollment, and protocols can be found at https://adni.loni.usc.edu.

### Cholesterol and Triglyceride measurements

Serum Cholesterol and Triglyceride levels were only assessed at baseline and mainly reported in mg/dL. Laboratory normal ranges were 0 - 199 mg/dL for Cholesterol and 0 - 149 mg/dL for Triglyceride. Defect and duplicated measurements were checked for each individual and removed based on the date and time of the reported result. Serum levels corresponding to 200 mg/dL for Cholesterol and 150 mg/dL for Triglyceride are largely recognized clinical cutoff values for hypercholesterolemia and hypertriglyceridemia, respectively. (19, 20) Owing to the larger use of mg/dL as a unit worldwide, values in mmol/L were converted to mg/dL:

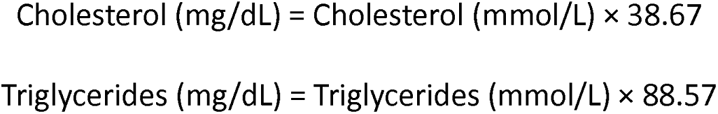

### Sleep and nighttime behavior disorders

The assessment of SNBD was based on the neuropsychiatric inventory questionnaire (NPI/NPI-Q) filled by study partners of included participants. (21, 22) The item related to sleep disorders in NPI/NPI-Q covered the following questions:

“Does the patient have difficulty sleeping (do not count as present if the patient simply gets up once or twice per night only to go to the bathroom and falls back asleep immediately)? Is he/she up at night? Does he/she wander at night, get dressed, or disturb your sleep? “

If this question was answered with yes, details on the following questions were then collected:

1. “Does the patient have difficulty falling asleep?”
2. “Does the patient get up during the night?”
3. “Does the patient wander, pace, or get involved in inappropriate activities at night?”
4. 4. “Does the patient awaken you during the night?”
5. “Does the patient wake up at night, dress, and plan to go out, thinking that it is morning and time to start the day?”
6. “Does the patient awaken too early in the morning, earlier than was his/her habit?”
7. “Does the patient sleep excessively during the day?”
8. “Does the patient have any other nighttime behaviors that bother you and we haven’t talked about?”

### Cognitive tests

Cognition was assessed based on the Alzheimer’s Disease Assessment Score with 13 items (ADAS_13_), Mini-Mental Status Examination (MMSE) total score, Clinical Dementia Rating (CDR) total score, CDR-sum of boxes (CDR-SB), and Functional Activities Questionnaire (FAQ) total score. Moreover, depression symptoms were reported based on the total Geriatric Depression Scale (GDS) score. People with severe depression were initially excluded from ADNI and therefore included participants are either non- or mildly depressed. The included cases were either healthy controls (HC), participants with mild cognitive impairment (MCI), or dementia.

### Body-mass index

Weight and Height data were checked to ensure the plausibility of the values and units. Weight at baseline was considered and converted to “Kilogram” when the unit of measurement was “Pounds”:

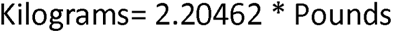

Similarly, height was converted to “Meters” when the unit of measurement was “Inches”:

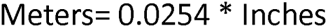

Body mass index (BMI) was calculated based on the formula weight (Kg) /Height (m). (23)

### Inclusion criteria

After excluding 142 participants without Cholesterol or Triglyceride measurements, 27 without complete NPI/NPI-Q, 21 missing total ADAS_13_ and one missing GDS score at baseline, nine missing baseline main diagnosis, and four missing complete demographic data (age), four missing weight or height at baseline and six have erroneous measurements or units, a total of 2,216 study participants were included in the analysis (fig. 1A). Among those 371 were lost to follow-up at 12 months (fig. 1B).

**Figure 1:**
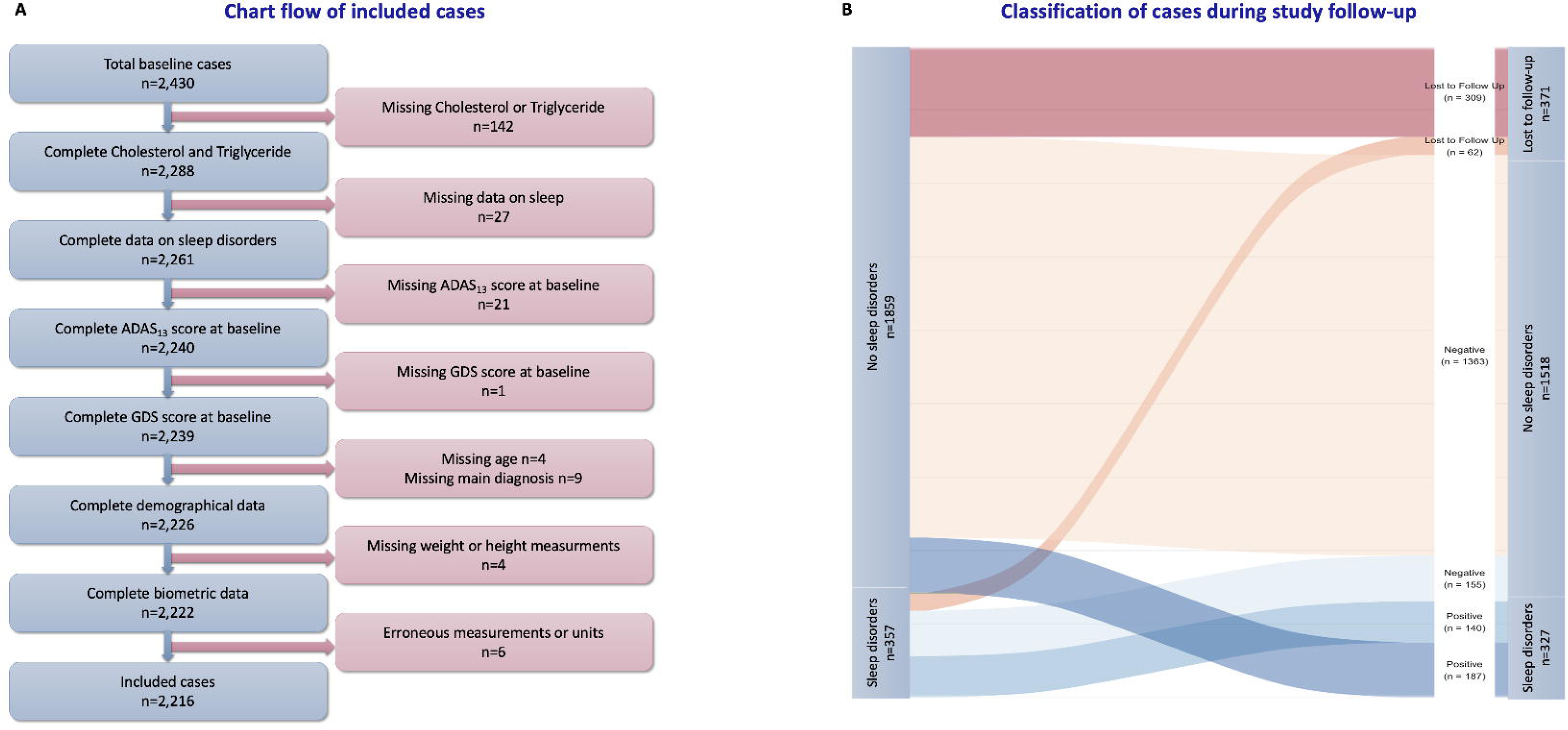
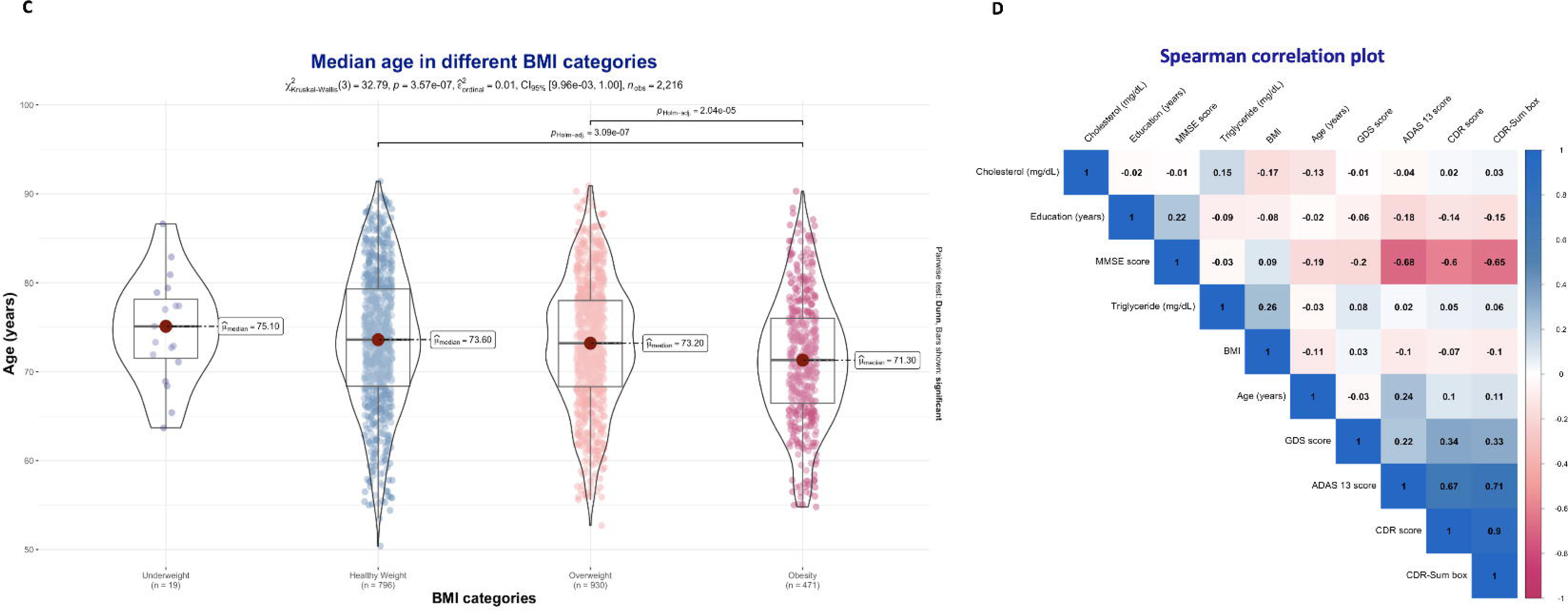
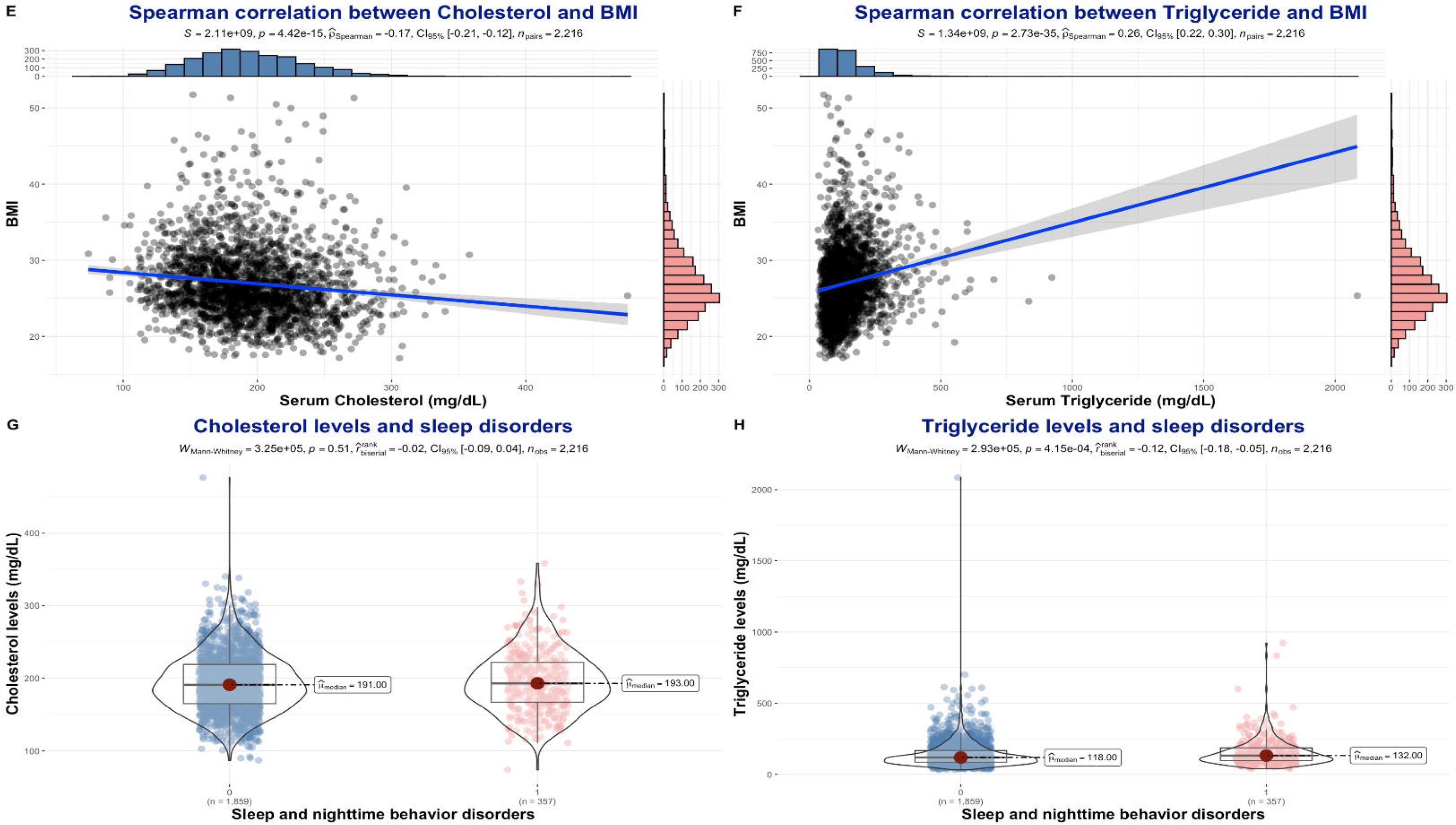
Characteristics of the study population Figure 1 A: Chart flow of included studies Figure 1 B: Classification of cases during study follow-up Figure 1 C: Median age across different BMI categories Figure 1 D: Spearman correlation plot Figure 1 E: Spearman correlation between Cholesterol and BMI Figure 1 F: Spearman correlation between Triglyceride and BMI Figure 1 G: Cholesterol levels and sleep disorders Figure 1 H: Triglyceride levels and sleep disorders

### Statistical analysis

The statistical analysis was performed by RStudio version 2024-04. Continuous data was reported as median (Inter-quartile range (IQR)) and count data as number (percentage (%)). Kruskal-Wallis rank sum test and Pearson’s Chi-squared test were performed to compare groups and for each analysis the *p*-value was reported. Spearman correlation between serum Cholesterol, Triglyceride, age, and scores of cognitive tests, was performed and correlation coefficients were reported. The association between SNBD and dyslipidemia at baseline was evaluated using logistic regression with SNBD as a dependent binary variable, and dyslipidemia as an independent binary variable. Models were adjusted for age, sex, racial profile, educational level, GDS total score, APOE ε4 status, main diagnosis related to cognitive status, and BMI, as following:

**Model 1:** crude logistic regression analysis,

**Model 2:** adjusted for age, sex, racial profile, educational level, cognition-related main diagnosis, geriatric depression scale total score, APOE ε4 status,

**Model 3:** model 2 + BMI

Incident cases of SNBD were calculated as new positive cases at 12 months of follow-up, amongst cases that were negative at baseline. Prediction analysis was based on logistic regression with SNBD at 12 months follow-up as a dependent binary variable, and dyslipidemia at baseline as an independent binary variable. The same confounding factors were adjusted for. For each model odds ratio (OR), 95% confidence interval (*CI*), and *p*-value were reported. The statistical significance level was set at 0.05.

## 3. Results

Among the 2,216 included cases, 1,045 (47%) were females, and 1,171 (53%) were males. The median age was 73 (IQR: 68, 78), and 786 (%) were HC, 1,060 (%) had MCI, and 370 (%) were diagnosed with dementia. The difference in median age between groups was statistically significant (72, 73, and 75 years, respectively, *p*-value <0.001).

At baseline, 357 (16%) study participants had sleep and nighttime behavior disorders, according to their study partner. At 12 months of follow-up, 327 (18%) participants had positive sleep disorder scores, among which 187 were incident cases (fig. 1B).

The median ADAS_13_ total score was 14 (IQR: 9, 22), the median GDS total score was 1.0 (IQR: 0, 2.0), and the median FAQ was 1.0 (IQR: 0, 5.0). Further characteristics of the included cases are presented in Table 1.

**Table 1:**
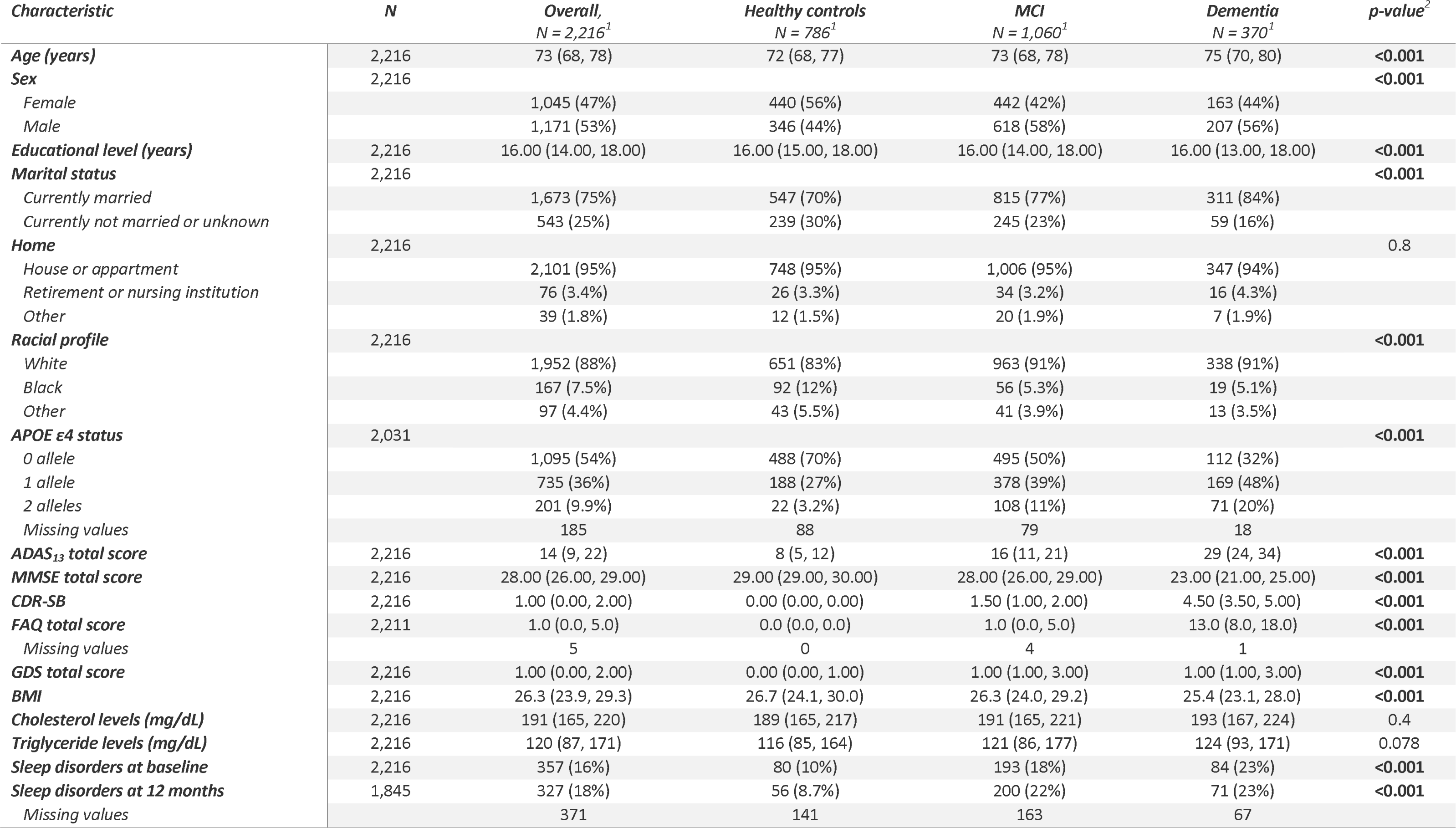
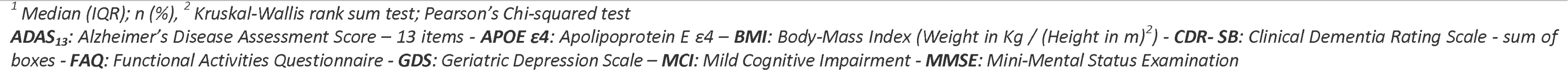
Characteristics of the study population and comparison between healthy controls, and those with MCI and dementia.

| *Characteristic* | *N* | *Overall,*<br>*N = 2,216<sup>1</sup>* | *Healthy controls*<br>*N = 786<sup>1</sup>* | *MCI*<br>*N = 1,060<sup>1</sup>* | *Dementia*<br>*N = 370<sup>1</sup>* | *p-value<sup>2</sup>* |
| --- | --- | --- | --- | --- | --- | --- |
| **Age (years)** | 2,216 | 73 (68, 78) | 72 (68, 77) | 73 (68, 78) | 75 (70, 80) | **<0.001** |
| **Sex** | 2,216 |  |  |  |  | **<0.001** |
| *Female* |  | 1,045 (47%) | 440 (56%) | 442 (42%) | 163 (44%) |  |
| *Male* |  | 1,171 (53%) | 346 (44%) | 618 (58%) | 207 (56%) |  |
| **Educational level (years)** | 2,216 | 16.00 (14.00, 18.00) | 16.00 (15.00, 18.00) | 16.00 (14.00, 18.00) | 16.00 (13.00, 18.00) | **<0.001** |
| **Marital status** | 2,216 |  |  |  |  | **<0.001** |
| *Currently married* |  | 1,673 (75%) | 547 (70%) | 815 (77%) | 311 (84%) |  |
| *Currently not married or unknown* |  | 543 (25%) | 239 (30%) | 245 (23%) | 59 (16%) |  |
| **Home** | 2,216 |  |  |  |  | 0.8 |
| *House or apartment* |  | 2,101 (95%) | 748 (95%) | 1,006 (95%) | 347 (94%) |  |
| *Retirement or nursing institution* |  | 76 (3.4%) | 26 (3.3%) | 34 (3.2%) | 16 (4.3%) |  |
| *Other* |  | 39 (1.8%) | 12 (1.5%) | 20 (1.9%) | 7 (1.9%) |  |
| **Racial profile** | 2,216 |  |  |  |  | **<0.001** |
| *White* |  | 1,952 (88%) | 651 (83%) | 963 (91%) | 338 (91%) |  |
| *Black* |  | 167 (7.5%) | 92 (12%) | 56 (5.3%) | 19 (5.1%) |  |
| *Other* |  | 97 (4.4%) | 43 (5.5%) | 41 (3.9%) | 13 (3.5%) |  |
| **APOE ε4 status** | 2,031 |  |  |  |  | **<0.001** |
| *0 allele* |  | 1,095 (54%) | 488 (70%) | 495 (50%) | 112 (32%) |  |
| *1 allele* |  | 735 (36%) | 188 (27%) | 378 (39%) | 169 (48%) |  |
| *2 alleles* |  | 201 (9.9%) | 22 (3.2%) | 108 (11%) | 71 (20%) |  |
| *Missing values* |  | 185 | 88 | 79 | 18 |  |
| **ADAS<sub>13</sub> total score** | 2,216 | 14 (9, 22) | 8 (5, 12) | 16 (11, 21) | 29 (24, 34) | **<0.001** |
| **MMSE total score** | 2,216 | 28.00 (26.00, 29.00) | 29.00 (29.00, 30.00) | 28.00 (26.00, 29.00) | 23.00 (21.00, 25.00) | **<0.001** |
| **CDR-SB** | 2,216 | 1.00 (0.00, 2.00) | 0.00 (0.00, 0.00) | 1.50 (1.00, 2.00) | 4.50 (3.50, 5.00) | **<0.001** |
| **FAQ total score** | 2,211 | 1.0 (0.0, 5.0) | 0.0 (0.0, 0.0) | 1.0 (0.0, 5.0) | 13.0 (8.0, 18.0) | **<0.001** |
| *Missing values* |  | 5 | 0 | 4 | 1 |  |
| **GDS total score** | 2,216 | 1.00 (0.00, 2.00) | 0.00 (0.00, 1.00) | 1.00 (1.00, 3.00) | 1.00 (1.00, 3.00) | **<0.001** |
| **BMI** | 2,216 | 26.3 (23.9, 29.3) | 26.7 (24.1, 30.0) | 26.3 (24.0, 29.2) | 25.4 (23.1, 28.0) | **<0.001** |
| **Cholesterol levels (mg/dL)** | 2,216 | 191 (165, 220) | 189 (165, 217) | 191 (165, 221) | 193 (167, 224) | 0.4 |
| **Triglyceride levels (mg/dL)** | 2,216 | 120 (87, 171) | 116 (85, 164) | 121 (86, 177) | 124 (93, 171) | 0.078 |
| **Sleep disorders at baseline** | 2,216 | 357 (16%) | 80 (10%) | 193 (18%) | 84 (23%) | **<0.001** |
| **Sleep disorders at 12 months** | 1,845 | 327 (18%) | 56 (8.7%) | 200 (22%) | 71 (23%) | **<0.001** |
| *Missing values* |  | 371 | 141 | 163 | 67 |  |

<sup>1</sup> Median (IQR); n (%), <sup>2</sup> Kruskal-Wallis rank sum test; Pearson's Chi-squared test
**ADAS<sub>13</sub>**: Alzheimer's Disease Assessment Score – 13 items - **APOE ε4**: Apolipoprotein E ε4 – **BMI**: Body-Mass Index (Weight in Kg / (Height in m)<sup>2</sup>) - **CDR-SB**: Clinical Dementia Rating Scale - sum of boxes - **FAQ**: Functional Activities Questionnaire - **GDS**: Geriatric Depression Scale – **MCI**: Mild Cognitive Impairment - **MMSE**: Mini-Mental Status Examination

Cholesterol levels ranged from 74 to 476 mg/dL, the median in the total population was 191 mg/dL (IQR: 165, 220), 189 mg/dL (IQR: 165, 217) in HC, 191 mg/dL (IQR: 165, 221) in the MCI group, and 193 mg/dL (IQR: 167, 224) in those with dementia. No statistically significant difference was found between diagnosis groups (*p*-value=0.4). The median Triglyceride level was 120 mg/dL (IQR: 87, 171) in the main population, and ranges between 32 and 2084 mg/dL. No statistically significant difference was found between diagnostic groups (116, 121, and 124 mg/dL, respectively, *p*-value=0.078). The median BMI of the main population was 26.3 (IQR: 23.9, 29.3), ranging between 17.14 and 51.75, with 19 (0.86%) classified as underweight, 796 (35.92%) as healthy weight, 930 (41.97%) as overweight, and 471 (21.25%) as obese. BMI showed a statistically significant difference between medians observed in different groups (26.7 in HC, 26.3 in MCI, and 25.4 in the dementia group, *p*-value <0.001).

Based on clinical cutoff values, hypercholesterolemia was diagnosed in 920 cases (41.52%), and hypertriglyceridemia in 725 cases (32.72%). Details on the differences between groups are presented in Table 2. BMI was significantly higher in the group without hypercholesterolemia than in those with (26.7 vs. 25.5, *p*-value <0.001). In contrast, BMI was higher in those with hypertriglyceridemia than in those without (25.7 vs. 27.5, *p*-value <0.001). BMI showed a decreasing tendency with age, cases with obesity were significantly younger than those with overweight and those with healthy weight (fig. 1C). Very weak correlations were found between hypercholesterolemia and hypertriglyceridemia with neurocognitive scores (fig. 1D). BMI was negatively correlated with serum Cholesterol levels, and positively correlated with serum Triglyceride levels (fig 1E and 1F).

**Table 2:**
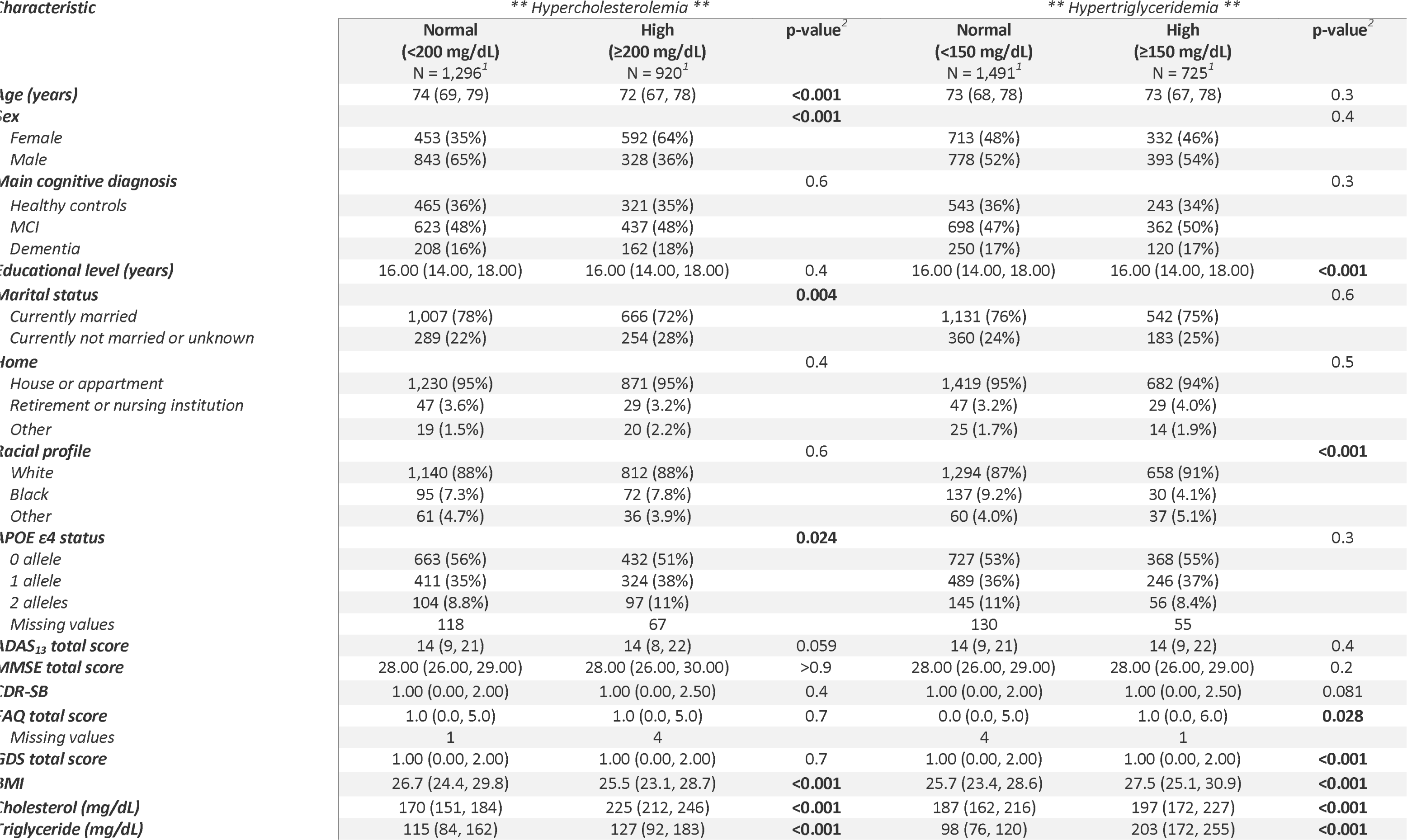
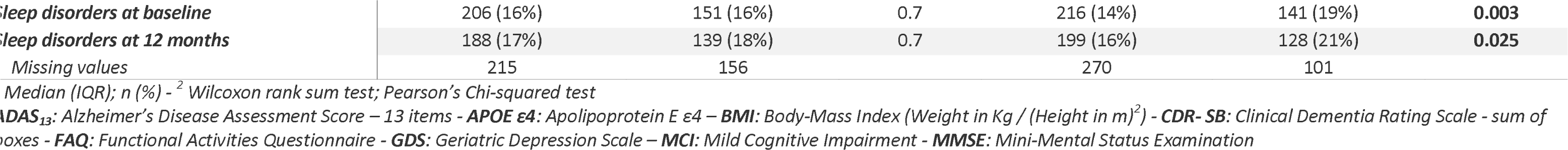
Comparison between study participants with and without dyslipidemia.

There was no significant difference in Cholesterol levels between those with SNBD at baseline and those without (16% vs. 16%, *p*-value=0.7). Similar results were also observed at 12 months (17% vs. 18%, *p-*value=0.7). Thus, there were more cases of SNBD in the hypertriglyceridemia group than in those with normal triglyceride (19% vs. 14%, *p*-value=0.003). Similarly, 21% of cases of SNBD at 12 months had hypertriglyceridemia and 16% had normal triglyceride levels at baseline (*p*-value=0.025). Cases with SNBD had higher serum Cholesterol levels than those without SNBD but the difference was not statistically significant (193 vs. 191 mg/dL, *p*-value=0.51) (fig. 1G). In contrast, cases with SNBD had significantly higher serum Triglyceride levels than those without (132 vs. 118 mg/dL, *p*-value <0.001) (fig. 1H).

At baseline, participants with hypercholesterolemia had 4% higher odds of SNBD but the results were not statistically significant (OR= 1.04, 95% *CI*: 0.83, 1.31, *p*-value=0.744). No statistical significance was observed after adjustment for confounding factors.

Those with hypertriglyceridemia levels had 43% higher odds of SNBD at baseline (OR= 1.43, 95% *CI*: 1.13, 1.80, *p*-value=0.003). The results remained statistically significant after adjustment for confounding factors (adj. OR=1.36, 95% *CI*: 1.06, 1.74, *p*-value=0.016) and after adding BMI to the adjusted model (BMI-adj. OR= 1.29, 95% *CI*: 1.00, 1.66, *p-* value=0.048). BMI and particularly obesity were significantly associated with higher odds of SNBD at baseline. No significant associations were found between hypercholesterolemia and hypertriglyceridemia with incident SNBD at 12 months of follow-up. Detailed results of the logistic regression were reported in Table 3.

**Table 3:**
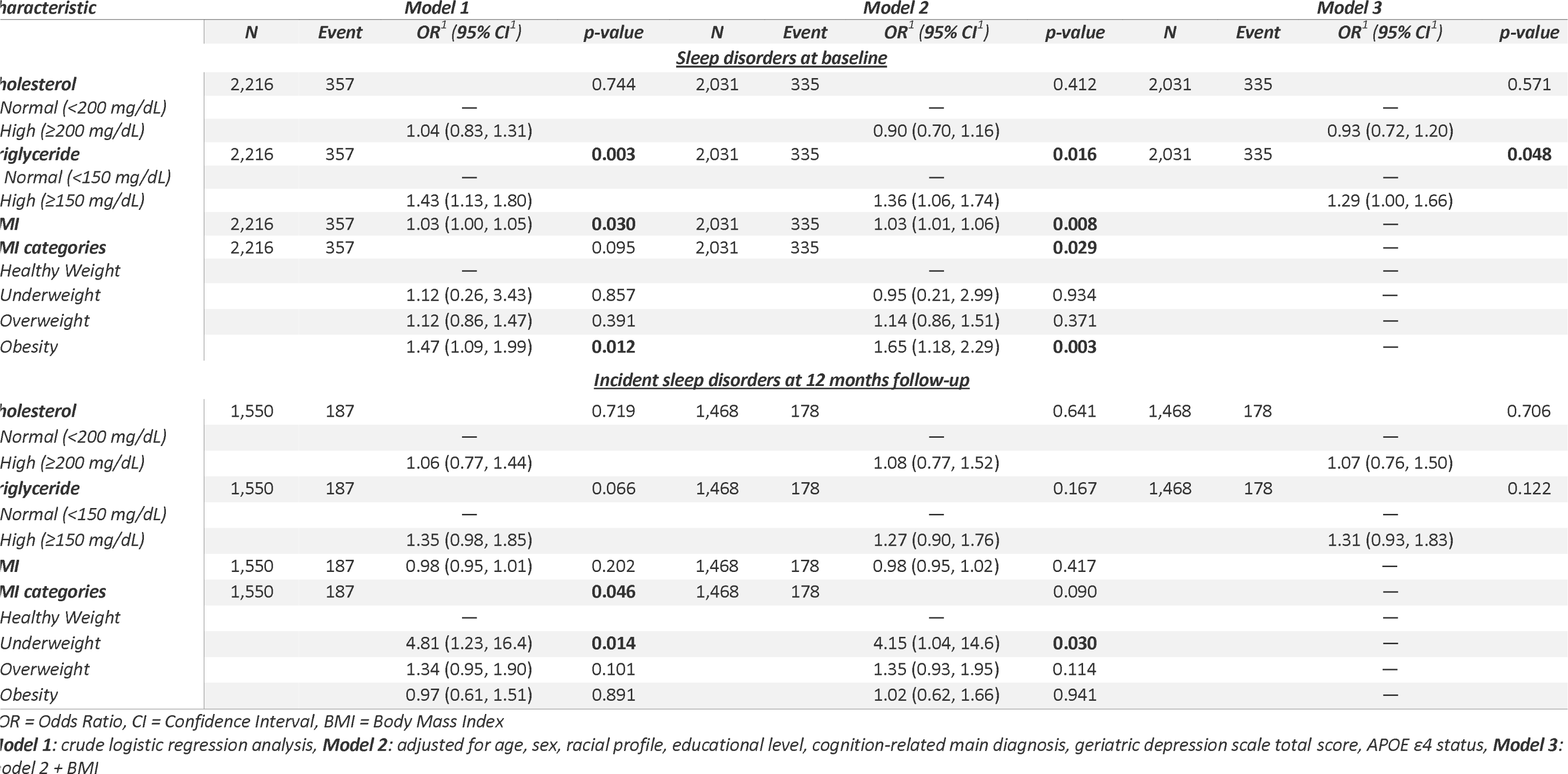
Association between sleep and nighttime behavior and dyslipidemia at baseline and 12 months follow-up.

| Characteristic | Model 1 |  |  |  | Model 2 |  |  |  | Model 3 |  |  |  |
| --- | --- | --- | --- | --- | --- | --- | --- | --- | --- | --- | --- | --- |
|  | N | Event | OR<sup>1</sup> (95% CI<sup>1</sup>) | p-value | N | Event | OR<sup>1</sup> (95% CI<sup>1</sup>) | p-value | N | Event | OR<sup>1</sup> (95% CI<sup>1</sup>) | p-value |
| **<u>Sleep disorders at baseline</u>** |  |  |  |  |  |  |  |  |  |  |  |  |
| **cholesterol** | 2,216 | 357 |  | 0.744 | 2,031 | 335 |  | 0.412 | 2,031 | 335 |  | 0.571 |
| Normal (<200 mg/dL) |  |  | — |  |  |  | — |  |  |  | — |  |
| High (≥200 mg/dL) |  |  | 1.04 (0.83, 1.31) |  |  |  | 0.90 (0.70, 1.16) |  |  |  | 0.93 (0.72, 1.20) |  |
| **triglyceride** | 2,216 | 357 |  | **0.003** | 2,031 | 335 |  | **0.016** | 2,031 | 335 |  | **0.048** |
| Normal (<150 mg/dL) |  |  | — |  |  |  | — |  |  |  | — |  |
| High (≥150 mg/dL) |  |  | 1.43 (1.13, 1.80) |  |  |  | 1.36 (1.06, 1.74) |  |  |  | 1.29 (1.00, 1.66) |  |
| **MI** | 2,216 | 357 | 1.03 (1.00, 1.05) | **0.030** | 2,031 | 335 | 1.03 (1.01, 1.06) | **0.008** |  |  | — |  |
| **MI categories** | 2,216 | 357 |  | 0.095 | 2,031 | 335 |  | **0.029** |  |  | — |  |
| Healthy Weight |  |  | — |  |  |  | — |  |  |  | — |  |
| Underweight |  |  | 1.12 (0.26, 3.43) | 0.857 |  |  | 0.95 (0.21, 2.99) | 0.934 |  |  | — |  |
| Overweight |  |  | 1.12 (0.86, 1.47) | 0.391 |  |  | 1.14 (0.86, 1.51) | 0.371 |  |  | — |  |
| Obesity |  |  | 1.47 (1.09, 1.99) | **0.012** |  |  | 1.65 (1.18, 2.29) | **0.003** |  |  | — |  |
| **<u>Incident sleep disorders at 12 months follow-up</u>** |  |  |  |  |  |  |  |  |  |  |  |  |
| **cholesterol** | 1,550 | 187 |  | 0.719 | 1,468 | 178 |  | 0.641 | 1,468 | 178 |  | 0.706 |
| Normal (<200 mg/dL) |  |  | — |  |  |  | — |  |  |  | — |  |
| High (≥200 mg/dL) |  |  | 1.06 (0.77, 1.44) |  |  |  | 1.08 (0.77, 1.52) |  |  |  | 1.07 (0.76, 1.50) |  |
| **triglyceride** | 1,550 | 187 |  | 0.066 | 1,468 | 178 |  | 0.167 | 1,468 | 178 |  | 0.122 |
| Normal (<150 mg/dL) |  |  | — |  |  |  | — |  |  |  | — |  |
| High (≥150 mg/dL) |  |  | 1.35 (0.98, 1.85) |  |  |  | 1.27 (0.90, 1.76) |  |  |  | 1.31 (0.93, 1.83) |  |
| **MI** | 1,550 | 187 | 0.98 (0.95, 1.01) | 0.202 | 1,468 | 178 | 0.98 (0.95, 1.02) | 0.417 |  |  | — |  |
| **MI categories** | 1,550 | 187 |  | **0.046** | 1,468 | 178 |  | 0.090 |  |  | — |  |
| Healthy Weight |  |  | — |  |  |  | — |  |  |  | — |  |
| Underweight |  |  | 4.81 (1.23, 16.4) | **0.014** |  |  | 4.15 (1.04, 14.6) | **0.030** |  |  | — |  |
| Overweight |  |  | 1.34 (0.95, 1.90) | 0.101 |  |  | 1.35 (0.93, 1.95) | 0.114 |  |  | — |  |
| Obesity |  |  | 0.97 (0.61, 1.51) | 0.891 |  |  | 1.02 (0.62, 1.66) | 0.941 |  |  | — |  |

OR = Odds Ratio, CI = Confidence Interval, BMI = Body Mass Index
**Model 1:** crude logistic regression analysis, **Model 2:** adjusted for age, sex, racial profile, educational level, cognition-related main diagnosis, geriatric depression scale total score, APOE ε4 status, **Model 3:** Model 2 + BMI

## 4. Discussion

This study explored the association between dyslipidemia based on pathological Cholesterol or Triglyceride levels at baseline, and sleep and nighttime behavior disorders reported by the study partner of included older participants at two time points.

The main outcome was the significant association between hypertriglyceridemia and sleep disorders in the cross-sectional analysis at baseline, even after adjusting for age, sex, racial profile, educational level, GDS total score, APOE ε4, cognition-related main diagnosis, and BMI.

### Sleep duration and dyslipidemia

Although insomnia is a well-recognized risk factor for cardiovascular and metabolic complications, (24, 25) the significant association between dyslipidemia and sleep disorders extends beyond sleep deprivation. Several population-based studies described a U-shaped association between sleep duration and serum lipid levels, where both short and long sleep durations were significantly associated with dyslipidemia. In people with longer sleep durations, high Triglyceride levels were commonly described. (26–28)

The novelty in the current study was the analysis of associations based on clinical cutoff values defining hypercholesterolemia or hypertriglyceridemia, and a binary outcome defining the existence or not of SNBD. Information on sleep hours was not part of the ADNI investigations, and the focus was SNBD as an outcome rather than sleep duration as an exposure.

### Sleep quality and dyslipidemia

In addition to the metabolic effect associated with the quantitative dimension of sleep, lipid levels might also be modulated by the subjective sleep quality. (14) Difficulty in maintaining sleep and excessive daytime sleepiness increased the odds of metabolic syndrome in the elderly, independently of obesity and snoring. (29) Moreover, the consumption of sleep medication, a biomarker of sleep disorder severity, showed a significant association with elevated low-density lipoprotein-cholesterol (LDL-C). (30) Neither LDL-C nor heigh-density lipoprotein-C (HDL-C) were reported in ADNI laboratory data.

### Sleep dysregulation and dyslipidemia

Night work, sleep debt, and social jetlag present further risk factors impairing sleep homeostasis and are associated with dyslipidemia and cardiovascular risks. (31, 32) Amongst 5,813 study participants from the Korean National Health and Nutrition Examination Survey (2013–2016), males exercising night work had 53% higher odds of being diagnosed with dyslipidemia. Compared to day-working male participants, male night workers who slept less than six hours and those who skipped meals had significantly higher odds of dyslipidemia. (33)

### Sleep apnea syndrome and dyslipidemia

The association between OSA and metabolic syndrome is well-described in the literature. (34, 35) Thus, independent of the OSA diagnosis, frequent snoring was also associated with dyslipidemia and predicted linearly higher levels of Triglyceride in a large population study. (36)

### Associations in younger patients

The focus of the current study was the elderly with and without cognitive decline. But, the sleep-lipid association was described in younger age groups as well. Higher Triglyceride levels were also reported in adolescents with longer sleep hours, (37) and accelerometry-based sleep clustering also showed that male adolescents with sleep irregularities had significantly higher Triglyceride levels. (38)

### Longitudinal studies

The second outcome was the absence of a significant association between dyslipidemia at baseline and incidental SNBD over a 12-month follow-up period.

While cross-sectional design is inadequate in inferring the causal relationship between sleep and dyslipidemia, longitudinal studies in older adults showed a bidirectional association between sleep duration and blood lipids. Total Cholesterol, LDL-C, HDL-C, and Triglycerides showed different temporal relationships with sleep duration. BMI and age were significant effect modifiers in this association. (39) In a longitudinal population-based cohort of healthy adults, short sleep duration increased the risk of metabolic syndrome, particularly hypertriglyceridemia by 9%. In comparison, long sleep duration decreased the risk of hypertriglyceridemia by 11%. (40)

### Mechanisms

The significant association between sleep disorders and higher Triglyceride levels might be explained by concomitant stress, frustration, and consequently eating irregularities. Stress and anxiety are associated with both sleep and eating disorders. (41) First, the association between sleep and stress is bidirectional; an increased emotional stress level might lead to sleep irregularities or insomnia, and sleep disorders might cause higher stress and frustration. (42) Sleep is crucial for emotion regulation and vice versa. (43, 44) Second, sleep disturbance interacts with hormone secretion and eating disorders. (45) Studies have shown that after sleep deprivation, people express eating and appetite dysregulation. (46, 47) Ghrelin, Leptin, and Adiponectin secretion patterns present a mediating effect on the association between sleep duration, and metabolic syndrome. (48, 49) Further, autoimmunity and neuroinflammation are relevant mechanisms involved in the homeostatic dysregulation associated with sleep disorders, and higher inflammatory biomarkers might further impair metabolic function and energy regulation. (50) Finally, a genetic predisposition, particularly Apolipoprotein genes, might infer the association between sleep and dyslipidemia. (51)

The sleep-lipid association was controversially discussed. Linear models seem insufficient to explain the association. (38) Moreover, published results were mainly different depending on the adjustment model used in the analysis. Noteworthy, adjusting for BMI and OSA led to the loss of the statistical significance of the association between sleep duration and quality on one side, and serum Triglyceride and hepatic Triglyceride content on another side. (16) It is known that BMI and OSA infer sleep quality and lipid levels and therefore might present a confounding effect on the path between sleep and dyslipidemia. This contradicts our results since associations remained statistically significant even after adjusting for BMI, and the questions on which the analysis of SNBD was based have mainly a behavioral aspect without considering respiratory symptoms.

## Limitations

Despite the interesting findings reported in this study, it is important to acknowledge some limitations.

The first limitation is related to the absence of information on LDL-C and HDL-C levels; both of which are relevant biomarkers of metabolic syndrome and dyslipidemia; but were not assessed in ADNI. Moreover, there was no clear information on whether blood measurements were performed during fasting. Most of which were collected between 10 am and 2 pm. Thus, a study on the association between sleep disturbances and Triglyceride levels in adolescents showed that the results were not affected by the fasting status of study participants and were statistically significant before and after stratifying by fasting during blood sampling. (38)

The second limitation is the absence of subjective data such as sleep duration or detailed polysomnographic measurements (Gold Standard). Furthermore, neither information on the objective description of the sleep quality reported by study participants, nor the presence or absence of obstructive sleep apnea were included. While those explorations are of high interest and might have enriched diagnostic methods and information used in the current study, they were largely explored in published data. The informant-based evaluation was favored, particularly because of its relevance in the elderly with cognitive impairment who might lack to some extent self-awareness and tend to over- or underestimate their sleep duration. The questionnaire was oriented toward sleep behavior disorders rather than breathing disorders and associated disturbances.

The third limitation is the lack of information on whether study participants were under lipid-regulating medications. Although this information could be relevant for evaluating the overall prevalence of dyslipidemia in the included sample, the main study objective was to investigate the association between pathological lipid levels at baseline as predicting variable and concomitant sleep disorders, independently of the potential medication effect.

## Conclusions

Hypertriglyceridemia, but not hypercholesterolemia, was associated with SNBD in the elderly at baseline. Moreover, none of the dyslipidemia forms did predict incidental cases of SNBD over a follow-up period of 12 months. This study emphasizes the importance of systematic screening of sleep disorders in older patients and its adapted management in mitigating metabolic risks and preventing related cardiovascular complications. Informant-based interviews are helpful and provide complementary information on sleep disorders in older individuals with cognitive impairment.

## Data Availability

All data produced are available online ADNI website

https://adni.loni.usc.edu

## Acknowledgments

“Data collection and sharing for the Alzheimer’s Disease Neuroimaging Initiative (ADNI) is funded by the National Institute on Aging (National Institutes of Health Grant U19 AG024904). The grantee organization is the Northern California Institute for Research and Education.

In the past, ADNI has also received funding from the National Institute of Biomedical Imaging and Bioengineering, the Canadian Institutes of Health Research, and private sector contributions through the Foundation for the National Institutes of Health (FNIH) including generous contributions from the following: AbbVie, Alzheimer’s Association; Alzheimer’s Drug Discovery Foundation; Araclon Biotech; BioClinica, Inc.; Biogen; Bristol-Myers Squibb Company; CereSpir, Inc.; Cogstate; Eisai Inc.; Elan Pharmaceuticals, Inc.; Eli Lilly and Company; EuroImmun; F. Hoffmann-La Roche Ltd and its affiliated company Genentech, Inc.; Fujirebio; GE Healthcare; IXICO Ltd.; Janssen Alzheimer Immunotherapy Research & Development, LLC.; Johnson & Johnson Pharmaceutical Research &Development LLC.; Lumosity; Lundbeck; Merck & Co., Inc.; Meso Scale Diagnostics, LLC.; NeuroRx Research; Neurotrack Technologies; Novartis Pharmaceuticals Corporation; Pfizer Inc.; Piramal Imaging; Servier; Takeda Pharmaceutical Company; and Transition Therapeutics.”

